# Effect of Electronic Nicotine Delivery Systems on Cigarette Abstinence in Smokers with no Plans to Quit: Exploratory Analysis of a Randomized Placebo-Controlled Trial

**DOI:** 10.1101/2021.06.22.21259359

**Authors:** Jonathan Foulds, Caroline O. Cobb, Miao-Shan Yen, Susan Veldheer, Phoebe Brosnan, Jessica Yingst, Shari Hrabovsky, Alexa A Lopez, Sophia I. Allen, Christopher Bullen, Xi Wang, Chris Sciamanna, Erin Hammett, Breianna L. Hummer, Courtney Lester, John P. Richie, Nadia Chowdhury, Jacob T. Graham, Le Kang, Shumei Sun, Thomas Eissenberg

## Abstract

**Introduction:** The extent to which use of electronic nicotine delivery systems (ENDS) for smoking reduction leads to cigarette abstinence in smokers with no plans to quit smoking is unclear. This exploratory analysis examined the effects of ENDS delivering different amounts of nicotine on cigarette abstinence up to 24-week follow-up, in comparison to placebo or a behavioral substitute.

**Methods:** This four-arm parallel-group, randomized placebo-controlled trial took place at two academic medical centers in USA (Penn State Hershey and Virginia Commonwealth University). Participants were current adult smokers (N=520) interested in reducing but not planning to quit. They received brief advice and were randomized to one of four 24-week conditions, receiving either an eGo-style ENDS paired with 0, 8 or 36 mg/ml nicotine liquid (double-blind) or a cigarette-shaped tube, as a cigarette substitute (CS). Self-reported daily cigarette consumption and exhaled carbon monoxide (CO) were measured at all study visits. Outcomes included intent-to-treat, self-reported 7-day cigarette abstinence, biochemically confirmed by exhaled CO at 24 weeks after randomization.

**Results:** At 24 weeks, significantly more participants in the 36 mg/ml condition (14/130, 10.8%) than in the 0 mg/ml condition (1/130, 0.8%) and the CS condition (4/130, 3.1%) were abstinent (relative risk = 14 [95% CI=1.9-104.9] and 3.5 [95% CI=1.2-10.4], respectively). The abstinence rate in the 8 mg/ml condition was 4.6% (6/130).

**Conclusions:** When smokers seeking to reduce smoking tried ENDS, few quit smoking in the short term. However, if smokers continued to use an ENDS with cigarette-like nicotine delivery, a greater proportion completely switched to ENDS, as compared with placebo or a cigarette substitute.

**IMPLICATIONS:** The extent to which use of electronic nicotine delivery systems (ENDS) for smoking reduction leads to cigarette abstinence in smokers with no plans to quit smoking was unclear. This randomized trial found that ENDS with nicotine delivery approaching that of a cigarette are more effective in helping ambivalent smokers to quit cigarette smoking.

## INTRODUCTION

Many smokers who try electronic nicotine delivery systems (ENDS) do so intending to reduce or quit smoking^1,2^ and some continue to “dual use”. The extent to which ENDS use for smoking reduction leads to increased cessation in smokers with no initial plans to quit smoking remains unclear.

An expert committee of National Academies of Science Engineering and Medicine ^3^ concluded that there is limited evidence that ENDS may be effective smoking cessation aids, and moderate evidence from randomized controlled trials (RCTs) that ENDS with nicotine are more effective than ENDS without nicotine. Since that report, some additional RCTs have found that ENDS increase smoking cessation rates^4-6^, but others did not^7-8^. Most RCTs of ENDS for smoking cessation to date have used ENDS with unknown or very low nicotine delivery, and many ENDS deliver very little nicotine relative to a cigarette^9-10^.

This report is based on exploratory analyses of a randomized placebo-controlled trial that was primarily designed to examine the effects of ENDS use on toxicant exposure in smokers. The methods and results for the primary toxicant outcome (4-(methylnitrosamino)-1-(3-pyridyl)-1-butanol, NNAL) have been reported elsewhere^11,12^. The primary outcome paper reported that participants assigned to 36 mg/ml nicotine ENDS reduced NNAL, exhaled carbon-monoxide (CO) and cigarette consumption^12^. Here we report on exploratory analyses that were planned prior to the data set being unblinded, and were designed to examine the effects of ENDS with differing nicotine delivery profiles^13^ on cigarette abstinence among current smokers with no plans to quit. We hypothesized “ECIG nicotine concentration-related reductions in combustible cigarette use” (SAP, p5), i.e. that the 36 mg/ml nicotine concentration ENDS would result in greater cigarette abstinence.

## METHODS

### Participants

Participants were 520 adult cigarette smokers of ≥10 cigarettes per day (CPD) who were interested in reducing their cigarette consumption by fifty percent but had no plans to quit within the next 6 months. For inclusion, volunteers were required to be aged 21-65, blow an exhaled CO of > 9 parts per million (ppm) at assessment and not have made any quit attempt or used a smoking cessation medication in the prior month. Participants could have used an ENDS product before, but not for more than 5 days out of the prior 28 days. Exclusions included any reported medical disorder/medication that may affect participant safety or biomarker data (e.g. pregnancy, unstable serious medical or psychiatric conditions, or weekly or greater use of illegal drugs). Full inclusion/exclusion criteria have been published^11,12^ and are available in the Supplement 1.

Recruitment occurred between July 22, 2015, and November 16, 2017, and follow-ups completed June 2018, at Penn State Medical Center in Hershey, Pennsylvania, (n=300), and Virginia Commonwealth University in Richmond, Virginia, (n=220), both in USA.

### Design

This was a four-arm, parallel-group, placebo-controlled randomized trial, with the three ENDS conditions administered double-blind^11,12^. Participants were randomized to one of four 24-week conditions and received either an eGo-style ENDS paired with 0, 8, or 36 mg/ml nicotine liquid, with tobacco or menthol flavor (participants chose flavor), or a cigarette-shaped plastic tube with no electronics or aerosol, for use as a cigarette substitute (CS). Study products were provided at no cost for up to 24 weeks. In previous laboratory studies, after 10 puffs over 5 minutes on the same ENDS device used in this trial, the 8 mg/ml and 36 mg/ml liquids gave a boost to blood nicotine concentrations of 6 ng/ml and 13 ng/ml^13^. The 36 mg/ml nicotine liquid used with this device therefore delivers a nicotine boost approaching that resulting from smoking a cigarette (around 15 ng/ml)^13^.

### Procedures

Participants were recruited via study advertisements including print, radio and online methods. They attended a screening visit and at the subsequent randomization visit (week 0), were asked to use their assigned product to help reduce their cigarette consumption by 50% through week 2 and then by 75% or more thereafter (relative to baseline). Instructions for participants were designed to be relatively simple and generic after they were informed how to use their study product. For example, at the randomization visit, all participants were told, “*As you learned during the consent process, you are being asked to reduce your traditional cigarette consumption throughout this study. Tomorrow, you will cut back on the number of cigarettes you smoke per day by half. This means that if you smoke 20 cigarettes per day, you will now smoke no more than 10 cigarettes per day. Remember the study product that we just discussed? You should use that study product to replace the cigarettes that you normally smoke*.”

At visit 4 (day 14), participants were told: *“During the next 14 days, you will be asked to reduce your cigarette consumption a bit more. Previously, we asked you to reduce your cigarette consumption by half. Now we are going to ask that you reduce your cigarette consumption by 75% compared to your baseline level. This means that if you smoked 20 cigarettes per day at the start of this study, you will now try to smoke only 5 cigarettes per day*.*”*

At visit 6 (day 56), participants were told: “*During the next 28 days, we want you to make sure you continue to attempt to achieve or maintain your 75% reduction in your cigarette consumption. We also want to encourage you to continue to use your study product to replace the cigarettes you have cut out*.”

Participants were provided with gift cards worth approximately $20-40 for each visit completed, up to a total of $400 for those completing all study procedures. These payments were intended to cover time and travel expenses and were not contingent on smoking reduction. Self-reported CPD and product use recorded via a paper daily diary were used to perform a 7-day timeline follow-back procedure at each of 8 visits (at weeks 1, 2, 4, 8, 12, 16, 20 and 24) during the intervention period. Exhaled carbon monoxide (CO) was measured at each visit, enabling validation of reported cigarette abstinence.

### Outcomes and statistical analysis

The original primary outcome of this trial was the carcinogen biomarker, NNAL, and the results on that outcome have been reported previously^12^. The trial also collected a number of additional measures and plans to conduct exploratory analyses on these were stated in the Statistical Analysis Plan (see SAP dated August 3, 2018 in Supplementary appendix 2, p16) prior to unblinded data analyses being initiated. Consistent with this SAP prior to unblinded analyses being undertaken, the cigarette abstinence-related outcomes are **(a)** intent-to-treat, self-reported 7-day point prevalence cigarette abstinence (PPA), biochemically confirmed by exhaled CO<10ppm (7-day PPA) for each visit up to 24 weeks after randomization (last visit of randomized phase of the trial), with those not attending visits counted as smoking. Additional outcomes included **(b)** self-reported 28 or more days of cigarette abstinence at week 24 (biochemically validated by exhaled CO<10ppm at weeks 20 and 24), **(c)** the number (%) of participants in each group who reported at least one full day without smoking a cigarette (no biochemical verification), from week 1 to week 24, and **(d)** the total number of days on which participants self-reported being abstinent from cigarettes from week 1 to week 24. The parent study was originally powered to detect effects on toxicant exposure (NNAL)^12^, rather than cigarette abstinence, but with 130 participants/group, there was 75% power to detect as significant (p<0.05, 2-tailed) a difference of 9% (e.g. 3% v 12%) between 2 groups, using Fisher’s exact test (post-hoc power calculation). Fisher’s exact tests were used to compare proportions abstinent between groups. Wilcoxon rank sum test were used to compare number of days of abstinence. 95% confidence intervals of differences were calculated.

The study was approved by the Institutional Review Boards at both institutions. The original study protocol is published^11^ and statistical analysis plan prior to unblinded analysis is available in Supplement 2. This study followed the Consolidated Standards for Reporting Trials (CONSORT) reporting guideline for randomized clinical trials.

## RESULTS

The main baseline characteristics of the four randomized groups are presented in Table 1, showing that they were well matched. The CONSORT diagram (Figure 1) shows that overall, 332 participants (63.8%) continued to attend through to 24 weeks with no significant between-group difference in dropout rates^12^.

**Table 1.**
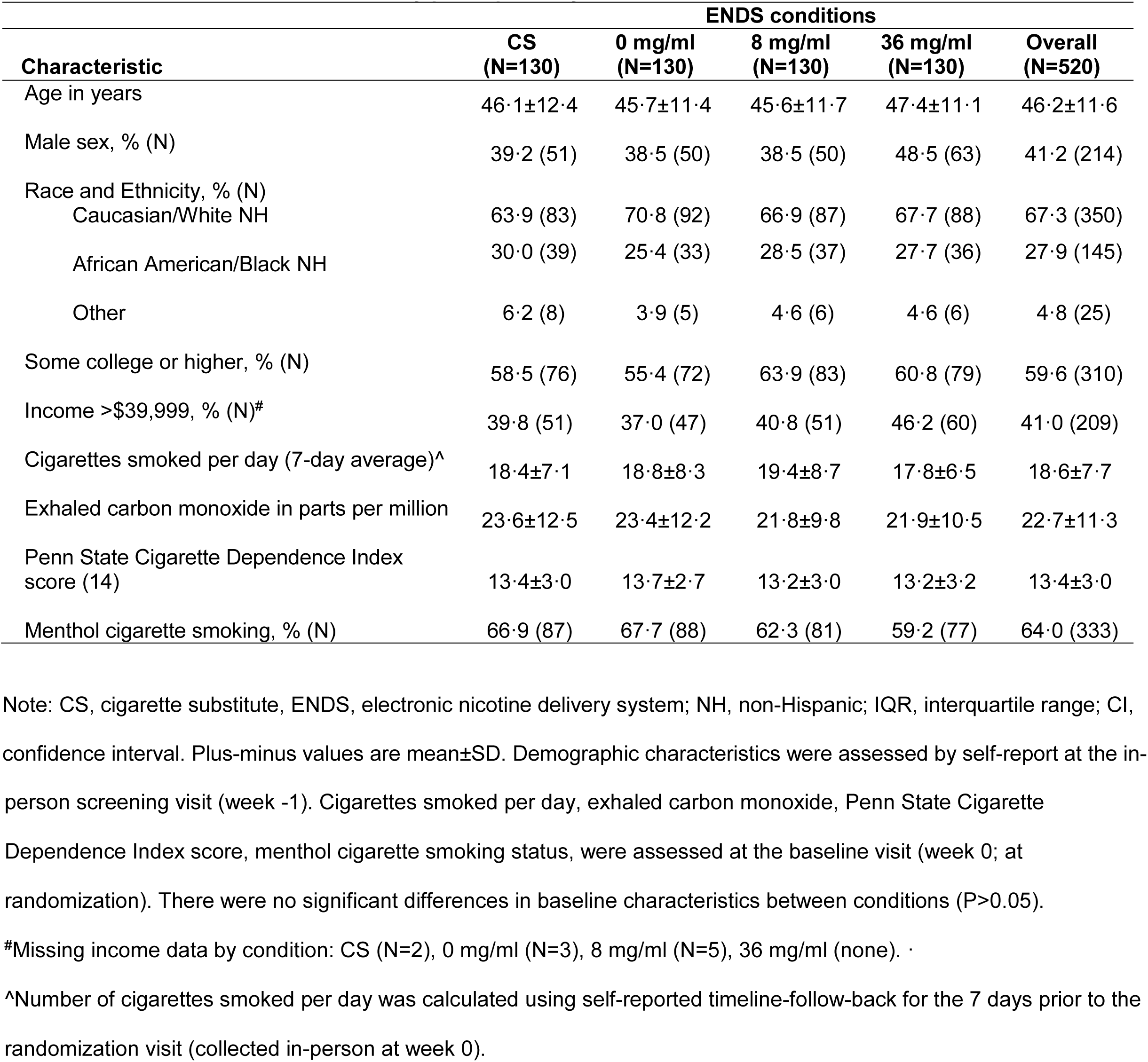
Baseline characteristics of study participants by condition and overall.

| **Characteristic** | **ENDS conditions** |  |  |  | **Overall**<br>**(N=520)** |
| --- | --- | --- | --- | --- | --- |
|  | **CS**<br>**(N=130)** | **0 mg/ml**<br>**(N=130)** | **8 mg/ml**<br>**(N=130)** | **36 mg/ml**<br>**(N=130)** |  |
| Age in years | 46.1±12.4 | 45.7±11.4 | 45.6±11.7 | 47.4±11.1 | 46.2±11.6 |
| Male sex, % (N) | 39.2 (51) | 38.5 (50) | 38.5 (50) | 48.5 (63) | 41.2 (214) |
| Race and Ethnicity, % (N) |  |  |  |  |  |
| Caucasian/White NH | 63.9 (83) | 70.8 (92) | 66.9 (87) | 67.7 (88) | 67.3 (350) |
| African American/Black NH | 30.0 (39) | 25.4 (33) | 28.5 (37) | 27.7 (36) | 27.9 (145) |
| Other | 6.2 (8) | 3.9 (5) | 4.6 (6) | 4.6 (6) | 4.8 (25) |
| Some college or higher, % (N) | 58.5 (76) | 55.4 (72) | 63.9 (83) | 60.8 (79) | 59.6 (310) |
| Income >\$39,999, % (N)<sup>#</sup> | 39.8 (51) | 37.0 (47) | 40.8 (51) | 46.2 (60) | 41.0 (209) |
| Cigarettes smoked per day (7-day average)<sup>^</sup> | 18.4±7.1 | 18.8±8.3 | 19.4±8.7 | 17.8±6.5 | 18.6±7.7 |
| Exhaled carbon monoxide in parts per million | 23.6±12.5 | 23.4±12.2 | 21.8±9.8 | 21.9±10.5 | 22.7±11.3 |
| Penn State Cigarette Dependence Index score (14) | 13.4±3.0 | 13.7±2.7 | 13.2±3.0 | 13.2±3.2 | 13.4±3.0 |
| Menthol cigarette smoking, % (N) | 66.9 (87) | 67.7 (88) | 62.3 (81) | 59.2 (77) | 64.0 (333) |

Note: CS, cigarette substitute, ENDS, electronic nicotine delivery system; NH, non-Hispanic; IQR, interquartile range; CI, confidence interval. Plus-minus values are mean±SD. Demographic characteristics were assessed by self-report at the in-
person screening visit (week -1). Cigarettes smoked per day, exhaled carbon monoxide, Penn State Cigarette Dependence Index score, menthol cigarette smoking status, were assessed at the baseline visit (week 0; at randomization). There were no significant differences in baseline characteristics between conditions ( $P>0.05$ ).

**Figure 1.**
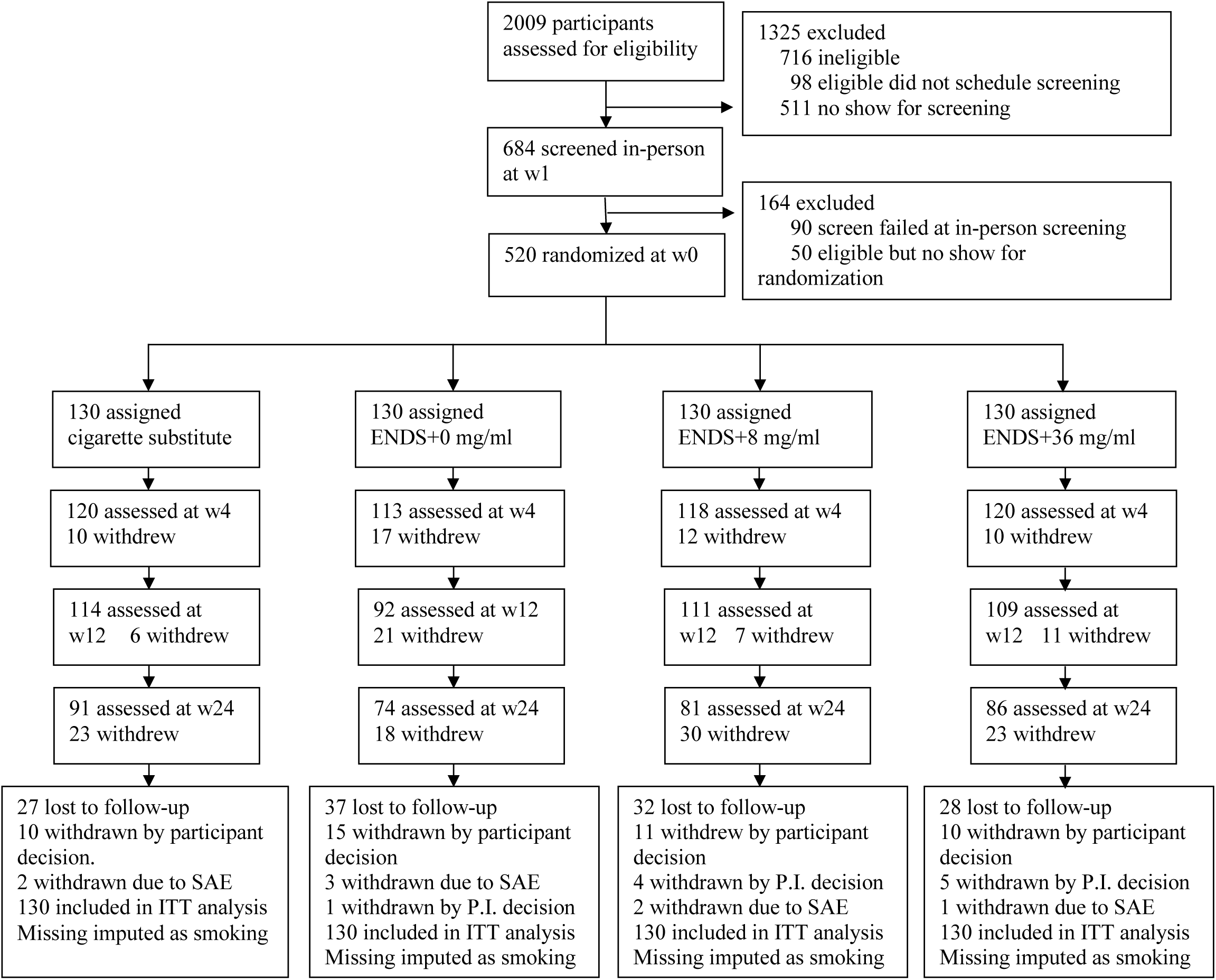
CONSORT flow diagram.

As shown in Figure 2, few participants in any group reported 7-day PPA within the first four weeks, when trying to reduce cigarette consumption by 50% and then 75%. However, over time more participants in the 36 mg/ml group achieved cigarette abstinence. At 24 weeks post-randomization, significantly more participants in the 36 mg/ml group than in the 0 mg/ml and CS groups were cigarette abstinent (Table 2). The mean exhaled CO among validated quitters in each group was <3ppm, as compared with 23 ppm at baseline. Participants randomized to 36 mg/ml were more likely to report at least one or more days of cigarette abstinence throughout the trial, at least 28 days abstinence at week 24, and more total days of cigarette abstinence (Table 2).

**Table 2.** Percentage of participants (n) who were abstinent from cigarettes at week 24, validated by exhaled carbon-monoxide (CO), (a) for the prior 7 days (b) for the prior 28+ days, (c) percentage (n) who reported at least one full day without cigarette smoking, and (d) mean number of total days without cigarette smoking in the 24 weeks of the trial (all n=130 per group, intent-to-treat, assuming smoking where data is missing) by group.

|  |  | GROUP |  |  |  |
| --- | --- | --- | --- | --- | --- |
| Outcome |  | Cig. Substitute (CS) | 0 mg/ml | 8 mg/ml | 36 mg/ml |
| **(a) 7-day point prevalence (CO<10) abstinence at week 24** | % (n) | 3.1 (4)# | 0.8 (1)* | 4.6 (6) | 10.8 (14) |
| | RR\$ (95% CI) | 3.50 [1.2,10.4] | 14.00 [1.9,104.9] | 2.33 [0.9,5.9] | |
| **(b) 28+ days abstinent with CO<10 at weeks 20 and 24~** | % (n) | 1.5 (2)# | 0.8 (1)* | 3.1 (4) | 7.7 (10) |
| | RR\$ (95% CI) | 5.50 [1.2,24.3] | 11.0 [1.4,84.0] | 2.75 [0.90,8.4] | |
| **(c) ≥1 day abstinent** | % (n) | 22.3 (29)* | 26.2 (34)# | 27.7 (36)# | 41.5 (54) |
| | RR\$ (95% CI) | 1.86 [1.3,2.7] | 1.59 [1.1,2.3] | 1.50 [1.1,2.1] | |
| **(d) Mean (SD) days no cigarette smoking** | Mean (SD) | 5.3 (18.5)^ | 4.7 (17.0)^ | 7.4 (23.1)^ | 15.6 (36.4) |
| | Difference of mean\$\$ (95% CI) | 10.29 [3.2,17.4] | 10.87 [3.9,17.8] | 8.20 [0.8,15.7] | |

\* $p < 0.006$ as compared to 36 mg/ml, Fisher's exact test.

**Figure 2.**
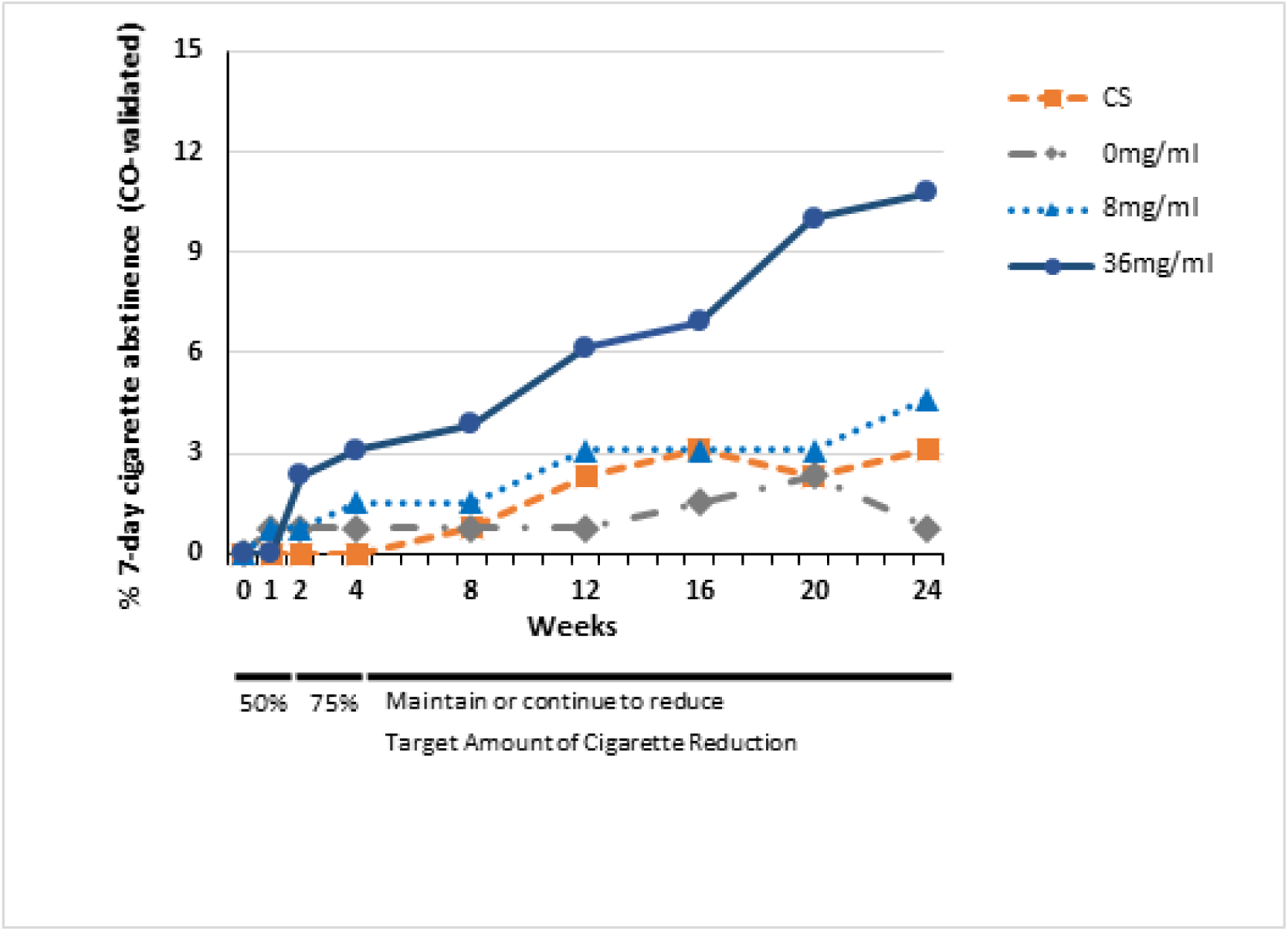
Percentage of participants in each randomized group reporting zero cigarette consumption in the prior 7 days at 8 follow-up visits, validated by exhaled CO<10ppm, at each visit. (0 mg/ml, 8 mg/ml or 36 mg/ml nicotine concentration in an Electronic Nicotine Delivery System or CS = Cigarette Substitute).

All 14 participants in the 36 mg/ml group who were abstinent at 24 weeks were using their assigned product when they first achieved abstinence (an average of 95 days earlier), and 12/14 of those abstainers (86%) were still using it at week 24.

Serious adverse events during the intervention period were spread evenly across the conditions (CS, 11; 0 mg/ml, 7; 8mg/ml, 5 and 36 mg/ml, 8) and none were judged to be related to study participation^12^.

## DISCUSSION

To our knowledge, this study is the first ENDS RCT in which a significant increase in cigarette abstinence compared with placebo ENDS and an alternative non-nicotine cigarette substitute, were observed. The abstinence rates were not high, and this is not surprising in daily smokers who expressed no plans to quit smoking at the start of the trial, who were encouraged to reduce smoking, and were given no explicit instructions to quit smoking during the trial. The extremely low quit rates in the placebo and CS groups (<1% and 3% at week 24) confirm that our study protocol did not stimulate spontaneous quitting.

While the ENDS with cigarette-like nicotine delivery (36 mg/ml) produced a marked increase in cigarette abstinence, there was no discernable beneficial effect on cigarette abstinence of the lower nicotine delivery ENDS. This result is consistent with dose-response effects on smoking cessation found with nicotine replacement therapies^15^. It suggests that policies that decrease the likelihood that smokers can identify and/or purchase ENDS that deliver nicotine as effectively as a combustible cigarette may have the unintended consequence of reducing the number of smokers who successfully quit smoking by switching from cigarettes to ENDS.

This study’s limitations include use of one ENDS device and no flavored liquids (other than tobacco and menthol), participant drop-out (36%), and a relatively short length of cigarette abstinence at 24 weeks. Strengths include randomized double-blind allocation to different liquid nicotine conditions, use of an ENDS device with known nicotine delivery profile, both placebo and non-ENDS control groups, and biochemical validation of self-reported abstinence.

## CONCLUSION

When smokers seeking to reduce smoking try ENDS, few quit smoking in the short term. However, if smokers continue to use an ENDS with cigarette-like nicotine delivery, a greater proportion completely switch to ENDS, as compared with placebo or a cigarette substitute. ENDS with nicotine delivery approaching that of a cigarette are more effective in enabling ambivalent cigarette smokers to quit smoking.

## Data Availability

With publication, requests for de-identified individual participant data and/or study documents (data dictionary, protocol, statistical analysis plan, measures/manuals/informed consent documentation) will be considered. The requestor must submit a 1-page abstract of their proposed research, including purpose, analytical plan, and dissemination plans. The Executive Leadership Committee will review the abstract and decide based on the individual merits. Review criteria and prioritization of projects include potential of the proposed work to advance public health, qualifications of the applicant, the potential for publication, the potential for future funding, and enhancing the scientific, geographic, and demographic diversity of the research portfolio. Following abstract approval, requestors must receive institutional ethics approval or confirmation of exempt status for the proposed research. An executed data use agreement must be completed prior to data distribution. Contact is through Dr. Caroline Cobb.

## ARTICLE INFORMATION

This research was supported by P50DA036105 and U54DA036105 from the National Institute on Drug Abuse of the National Institutes of Health and the Center for Tobacco Products of the U.S. Food and Drug Administration. Data collection was supported by UL1TR002649 at Virginia Commonwealth University and by UL1TR002014 at Penn State University from the National Center for Advancing Translational Sciences of the National Institutes of Health. Funding sources had no other role other than financial support. The content is solely the responsibility of the authors and does not necessarily represent the official views of the National Institutes of Health or the Food and Drug Administration.

## Notes

### Competing Interest Statement

JF reports a research grant, receipt of study medication, personal fees and non-financial support from Pfizer Inc., outside the submitted work. He has also purchased ENDS products for use in clinical trials. CB has previously undertaken trials of e-cigarettes for smoking cessation, requiring purchase of ENDS products and nicotine patches, outside the submitted work. None of the above parties had any role in the design, conduct, analysis or interpretation of the trial findings, or writing of the resulting publication. TE is a paid consultant in litigation against the tobacco industry and also the electronic cigarette industry and is named on one patent for a device that measures the puffing behavior of electronic cigarette users and on another patent for a smartphone app that determines electronic cigarette device and liquid characteristics.

### Clinical Trial

NCT02342795

### Author Declarations

Approved by IRBs at both Penn State University and Virginia Commonwealth University.

