## Supplement 1 for "Effect of Electronic Nicotine Delivery Systems on Cigarette Abstinence in Smokers with no Plans to Quit: Exploratory Analysis of a Randomized Placebo-Controlled Trial"

### Acknowledgements

#### Penn State University- College of Medicine

Jonathan Foulds, PhD

Erin Hammett, MS

Sharilee Hrabovsky, DEd

Breianna L. Hummer, MS

Courtney Lester, BS

John P. Richie, Jr., PhD

Susan Veldheer, DEd

Jessica M. Yingst, DrPH

Jennifer Modesto, MS

Richard Sargent, BA

Alyse Fazzi, PharmD

Sophia I. Allen, PhD

Lisa Reinhart, MS

Christopher Sciamanna, MD

Xi Wang MS

#### Virginia Commonwealth University

Thomas Eissenberg, PhD

Caroline O. Cobb, PhD

Phoebe Brosnan, MA

Nadia Chowdhury, BS

Parker Webster, BSN

Tyler Brunet, MS

Jacob T. Graham, BS

Alexa A. Lopez, PhD

Miao-Shan Yen, MS

Le Kang, PhD

Mishaal Khan, BS

Nancy Sey, BS

Sam Patel, BS

Matthew Miera, BS

Alyssa K. Rudy, MS

Thokozeni Lipato, MD

Shumei Sun, PhD

Matthew Halquist, PhD

Alison Montpetit, PhD

Susan Teasley

Jennifer Economy

**The University of Auckland**

Christopher Bullen, PhD

**Wake Forest University**

Eric Donny, PhD

**CSTP-Randomized Control Trial Methods Workgroup**

**Virginia Commonwealth University**

Thomas Eissenberg, PhD (Principal Investigator, VCU)

Caroline O. Cobb, PhD

Phoebe Brosnan, MA

Nadia Chowdhury, BS

Jacob T. Graham, BS

Alexa A. Lopez, PhD

Miao-Shan Yen, MS

Le Kang, PhD

Shumei Sun, PhD

Thokozeni Lipato, MD

**Penn State University-College of Medicine**

Jonathan Foulds, PhD (Principal Investigator, PSU)

Erin Hammett, MS

Sharilee Hrabovsky, DEd

Breianna L. Hummer, MS

Courtney Lester, BS

John P. Richie, Jr., PhD

Susan Veldheer, DEd

Jessica M. Yingst, DrPH

Christopher Sciamanna, MD

Sophia I. Allen, PhD

Xi Wang, MS

**The University of Auckland**

Christopher Bullen, PhD

**Data Safety Monitoring Board**

Dace Svikis, PhD (Chair), Professor, Departments of Psychology, Psychiatry, and Obstetrics/Gynecology, Virginia Commonwealth University

Leo Dunn, MD, Professor, Department of Obstetrics and Gynecology, Virginia Commonwealth University

Adam Sima, PhD, Associate Professor, Department of Biostatistics, Virginia Commonwealth University

Erika F.H. Saunders, MD, Associate Professor, Chair, Department of Psychiatry, Penn State College of Medicine

### Supplementary Methods

#### Contributions of authors

Dr. Foulds wrote the first draft of the paper and circulated to all authors for comments. Ms. Yen and Ms. Wang conducted all analyses in consultation with Dr. Foulds and other authors. All authors participated in designing the study, collecting the data, and/or analyzing and interpreting the data. All authors share in the decision to publish the paper and in the responsibility for the manuscript as submitted. Members of the CSTP-Randomized Control Trial Methods Workgroup contributed to designing the study and/or collecting/analyzing the data.

#### Study Protocol and Statistical Analysis Plan

A complete study protocol and statistical analysis plan is available in Supplementary Appendix B. A published version of the study protocol and original statistical analysis plan is publically available.[1]

#### Study products and blinding

Electronic nicotine delivery system (ENDS) liquid (0, 8, and 36 mg/ml) was purchased from an ENDS retailer located in Richmond, VA and had nicotine concentration verified by an independent laboratory prior to use (BLQ for 0 mg/ml; +/-1 mg/ml for 8 mg/ml; +/-2 mg/ml for 36 mg/ml). Following nicotine concentration verification, ENDS liquid was split between sites and shipped as needed for dispensation.

Cartomizer preparation and dispensation procedures were identical between sites and utilized paper-based logs as well as electronic records to ensure blinding as well as accurate product assignment. Administrative staff with no participant contact prepared all cartomizers for dispensation (1 mL of liquid per cartomizer). All filled cartomizers were stored with the cartomizer mouth-end upright in child-proof plastic vials (7 cartomizers per vial). Filled cartomizers/vials were discarded after 27 days to ensure cartomizers more than 4 weeks old were not given to participants. This procedure was used to as a quality control measure considering some study visits were approximately 4 weeks apart. When dispensed, child-proof plastic bottles (not cartomizers) were labeled with an adhesive sticker by administrative staff that indicated participant ID, liquid flavor, visit number, and date of cartomizer expiration.

Following randomization to the cigarette substitute condition, researchers reviewed and provided participants with a copy of their study product manual along with two cigarette substitutes. Manual instructions also included the following text: “For best results - Practice using your cigarette substitute (including trying different airflow settings) to find a method that works best for you.”

Following randomization to an ENDS condition, researchers reviewed and provided participants with a copy of their study product manual along with two pre-charged ENDS batteries, a charger, and carrying case. The ENDS manual provided detailed information on what participants were given, how to set up the ENDS (i.e., attach/replace the cartomizer, use the button to activate the heating element, safely store their study product), how to maintain their daily tobacco use diary, and some potential ENDS related side effects. Manual instructions also included the following text: "For best results - Practice using your ECIG to find a method that works best for you, consistent with local rules and regulations regarding clean indoor air." Following manual review, researchers then asked participants to sample two cartomizers corresponding to tobacco and menthol flavor of their assigned condition (i.e., liquid nicotine concentration was consistent with condition assigned). Participants were informed that they would receive the selected ENDS flavor for the duration of the study. Following sampling and flavor selection, researchers retrieved the participant's full supply of cartomizers for that visit.

Of note all participants had the opportunity to experience the alternate study product to which they were randomized at the 24-week visit (end of the intervention period). Individuals initially randomized to an ENDS condition received two cigarette substitutes and a study product manual at week 24. Individuals initially randomized to a cigarette substitute condition were provided with 21 cartomizers (always at 0 mg/ml; flavor consistent with cigarette menthol preference), 1 ENDS battery/charger/carrying case, and a study product manual.

Each ENDS+liquid concentration was tested in a clinical laboratory study using the same ENDS, cartomizer, and liquid with identical characteristics.[1] When experienced END users were asked to take 10 puffs, 8 mg/ml liquid resulted in a boost of 8.2 ng/ml (SD=7.8), and 36 mg/ml liquid resulted in a boost of 17.9 ng/ml (SD=17.2). When ENDS-naive cigarette smokers asked to take 10 puffs, 8 mg/ml liquid did not result in a significant increase in plasma nicotine relative to baseline, and 36 mg/ml liquid resulted in a boost of 6.8 ng/ml (SD=7.1).[2]

**Detailed Inclusion and Exclusion Criteria** (as described in Lopez et al, 2016)[1]

**INCLUSION**: For inclusion in the study, participants must

- be between the ages of 21–65
- report smoking >9 regular filtered cigarettes or machine-rolled cigarettes with a filter for at least 1 year and present with an expired air CO measurement of >9 parts per million at baseline.
- have made no serious cigarette smoking quit attempt in the prior 1 month. This criterion includes the use with the intent to quit cigarette smoking of any FDA-approved smoking cessation medication (varenicline; bupropion used specifically as a quitting aid; nicotine patch, gum, lozenge, inhaler, or nasal spray) in the past 1 month.

- not be planning to quit smoking in the next 6 months, they must report that they are interested in reducing their cigarette consumption by at least half in the next 6 months
- be willing to attend visits weekly and monthly over a 9-month period (i.e., not planning to move, take an extended vacation, undergo surgeries).
- be able to read and write in English and be able to understand and give informed consent.

**EXCLUSION**: Participants were excluded from the study if they

- were pregnant and/or nursing

- had any unstable or significant medical condition in the past 12 months that might lead to study exclusion (e.g., recent heart attack or some other heart conditions, stroke, severe angina including high blood pressure if systolic >159 or diastolic >99 observed during screening).

- had other health indicators for exclusion included immune system disorders, respiratory diseases (e.g., exacerbations of asthma or COPD, require oxygen, require oral prednisone), kidney (e.g., dialysis) or liver diseases (e.g., cirrhosis), or any medical disorder/medication that may affect participant safety or biomarker data.

- used any non-cigarette nicotine delivery product (e.g., pipe, cigar, dip, chew, snus, hookah, ECIGs, strips, sticks) in the past 7 days at the initial baseline assessment.

- had used an ECIG for 5 or more days in the past 28 days,

- had used marijuana or other illegal drugs daily/almost daily, or weekly in the past 3 months,

- use hand-rolled roll your own cigarettes

- had uncontrolled mental illness or substance abuse or inpatient treatment for these in the past 6 months,

- had history of difficulty providing or unwilling to provide blood samples (e.g., fainting, poor veins, anxiety)

- had planned surgery requiring general anesthesia in the past 6 weeks,

- had another member of household participated or currently participating in the study.

- had any known allergy to propylene glycol or vegetable glycerin

### RANDOMIZATION AND MASKING

The study statistician (Ms. Yen) prepared site-specific randomization lists using the sample function in R software (blocks of 8). These lists were uploaded onto a study-specific website that interfaced with the data collection/management system (REDCap). Only unblinded researchers at each site with no participant contact accessed their list to prepare cartomizers for dispensing. Participants were randomized in-person and in real-time using an electronic function within REDCap that revealed allocation to either an ENDS or CS condition at the time of randomization. Participants and researchers who enrolled participants and who collected outcome data were masked to which liquid nicotine concentration was assigned among individuals in ENDS conditions, and all cartomizers were identical except for the liquid placed within them. Masking success was not evaluated systematically. Those analyzing data were not masked to condition assignment.

#### Definition of cigarette abstinence.

For all calculations of cigarette abstinence, if a participant did not attend a visit to report abstinence or provide a sample of exhaled carbon-monoxide (CO), they were assumed to be smoking since the last visit when they reported abstinence. The definitions of abstinence in quotes below were taken from the Statistical Analysis Plan finalized 08/03/2018, (p16) prior to any unblinded analyses.

(a) The main definition of abstinence used in the Statistical Analysis Plan was "*Cigarette smoking abstinence (defined as 0 cigarettes smoked via 7-day average and expired air CO <10 at the same visit; SRNT Subcommittee, 2002)*." [3] For the **primary outcome analysis this was operationalized as self-reported 7-day cigarette abstinence validated by an exhaled CO <10 ppm (7-day PPA) at week 24.**

Additional definitions of abstinence were:

(b) "*28-day point prevalence cigarette smoking abstinence*". This was operationalized as **self-reported 7-day cigarette abstinence validated by an exhaled CO <10 ppm at week 24 and week 20, with no reported cigarettes in between**. In practice this represents 35 days of cigarette abstinence (28+7) but we refer to it as 28+ days as there was variability in appointment dates between visits.

"*24-hour point prevalence cigarette smoking abstinence*" was calculated in 2 ways using participant self- reports:

(c) the number (%) of participants in each group who reported **at least one full day without smoking a cigarette** (no biochemical verification), from week 1 to week 24, and

(d) the **total number of days on which participants self-reported being abstinent from cigarettes from week 1 to week 24** (no biochemical verification).

**Table S1 Number of randomized participants (%) who completed all 10 visits (2 pre-randomization, 8 post-randomization) to week 24.**

| | CS | 0 mg/ml | 8 mg/ml | 36 mg/ml | Total |
|---|---|---|---|---|---|
| **#completed/randomized** | 91/130 | 74/130 | 81/130 | 86/130 | 332/520 |
| **% completed** | 70.0 | 57.0 | 62.3 | 66.2 | 63.8 |

**Table S2. Study product use percentages using intent-to-treat method (with assumption that those not attending/providing data were not using study product)**

| Percent using study product, %, (n) | CS | 0 mg/ml | 8 mg/ml | 36 mg/ml |
|---|---|---|---|---|
| **Week 1** | 86·9 (113) | 85·4 (111) | 87·7 (114) | 89·3 (116) |
| **Week 2** | 77·7 (101) | 75·4 (98) | 84·6 (110) | 84·6 (110) |
| **Week 4** | 71·5 (93) | 66·9 (87) | 75·4 (98) | 81·5 (106) |
| **Week 8** | 44·6 (58) | 54·6 (71) | 56·9 (74) | 60·8 (79) |
| **Week 12** | 41·5 (54) | 42·3 (55) | 52·3 (68) | 55·4 (72) |
| **Week 16** | 34·6 (45) | 43·1(56) | 43·9 (57) | 53·9 (70) |
| **Week 20** | 36·2 (47) | 35·4 (46) | 40·8 (53) | 48·5 (63) |
| **Week 24** | 33·1 (43) | 36·2 (47) | 37·7 (49) | 47·7 (62) |

Note: CS, cigarette substitute. Participants with missing study product usage logs were assumed no use of study product. Denominator for calculating the percent using is always 130 for every condition.

**Table S3. Percentage (n) of participants in each group reporting smoking zero cigarettes in the prior 7 days, validated by exhaled CO <10ppm at each visit using intent-to-treat method (with assumption that those not attending/providing data were smoking)**

| Percent (n) cigarette abstinent | CS | 0 mg/ml | 8 mg/ml | 36 mg/ml |
|---|---|---|---|---|
| **Week 1** | 0 | 0.77 (1) | 0.77 (1) | 0 |
| **Week 2** | 0 | 0.77 (1) | 0.77 (1) | 2.31 (3) |
| **Week 4** | 0 | 0.77 (1) | 1.54 (2) | 3.08 (4) |
| **Week 8** | 0.77 (1) | 0.77 (1) | 1.54 (2) | 3.85 (5) |
| **Week 12** | 2.31 (3) | 0.77 (1) | 3.08 (4) | 6.15 (8) |
| **Week 16** | 3.08 (4) | 1.54 (2) | 3.08 (4) | 6.92 (9) |
| **Week 20** | 2.31 (3) | 2.31 (3) | 3.08 (4) | 10.00 (13) |
| **Week 24** | 3.08 (4) | 0.77 (1) | 4.63 (6) | 10.77 (14) |

Note: CS, cigarette substitute. Participants with missing data were assumed to be smoking. Denominator for calculating the percent using is always 130 for every condition.

**Table S4 Number and percentage of week 24 abstainers who were using their assigned study product when they first achieved 7-day abstinence.**

| | CS | 0 mg/ml | 8 mg/ml | 36 mg/ml | Total |
|---|---|---|---|---|---|
| N using/ n abstinent | 0/4 | 1/1 | 5/6 | 14/14 | 20/25 |
| % of week 24 abstainers | 0 | 100 | 83.3 | 100 | 63.8 |

**Table S5 Number and percentage of week 24 abstainers who were using their assigned study product at week 24.**

| | CS | 0 mg/ml | 8 mg/ml | 36 mg/ml | Total |
|---|---|---|---|---|---|
| N using/ n abstinent | 0/4 | 0/1 | 5/6 | 12/14 | 17/25 |
| % of week 24 abstainers | 0 | 0 | 83.3 | 85.7 | 68 |

**Table S6. Count of serious and severe adverse events**

| | | Intervention (W1-W24) | | | |
|---|---|---|---|---|---|
| | Overall | CS | 0 mg/ml | 8 mg/ml | 36 mg/ml |
| **N Serious Adverse Events** | 31 | 11 | 7 | 5 | 8 |
| **N Serious Adverse Events (possibly/probably/definitely related)*** | 0 | 0 | 0 | 0 | 0 |
| **N Severe AEs** | 71 | 19 | 21 | 14 | 17 |
| **N Severe Adverse Events (possibly/probably/definitely related)** | 4 | 0 | 1 | 2 | 1 |

Note: AE, adverse event; W, week. Multiple symptoms related to the same event in the same participant are only counted once. Study period when SAE noted may represent when symptoms first emerged (i.e. initial AE), not when the serious and/or severe event was documented. Serious AEs may have been designated as either mild/moderate/severe/life-threatening/fatal in terms of severity. Severe AEs include those coded as severe/life-threatening/fatal. * All serious AEs are unrelated or unlikely (remote)

**Non-study product use.**

As use of more than one nicotine product and other substances is very common among smokers, participants were included in the study (randomized per protocol) if they had used another nicotine product or substance in the prior 30 days (but not the prior 7 days), at baseline. Overall, 31 participants (6%) stated that they had used another tobacco product in the prior 30 days at baseline. Participants were encouraged to avoid other product use (i.e. non-randomized study products or cigarettes) throughout the trial, but they were encouraged to report such use and remain in the study. Occasional, very low rates of non-study product use were reported throughout the trial. With regards to the primary/secondary outcomes at week 24, at week 24, 4 participants who met criteria for cigarette abstinence stated that they had used a non-study product in the prior 7 days (all randomized to 36 mg/ml). 3 had used a non-study ENDS and one had used marijuana on 1 day. None of these participants had used a non-study product in the prior 7 days at the week 20 visit. At week 20, one participant who was cigarette abstinent during the previous 7 days at that visit reported using a non-study product. That participant had been randomized to the CS group, and reported using an ENDS during the prior 7 days at that visit. None of the participants who met the protocol-defined criteria for CO-verified cigarette abstinence at week 24 reported using smoked tobacco products in the prior 28-35 days (i.e. they did not report use of cigarettes, cigars, hookah etc).
