## Supplementary Appendix 2 for "Effect of Electronic Nicotine Delivery Systems on Cigarette Abstinence in Smokers with no Plans to Quit: Exploratory Analysis of a Randomized Placebo-Controlled Trial"

### Statistical Analysis Plan -

### Randomized Controlled Trial Methods for Novel Tobacco Products Evaluation

Version: 2

Caroline Cobb

Miao-Shan Yen

Le Kang

Shumei Sun

Thomas Eissenberg

Jonathan Foulds

Date: 8/3/2018

### Section 1: Administrative Information

**1.1. Title**

Randomized Controlled Trial Methods for Novel Tobacco Products Evaluation

**1.2. Trial Registration**

ClinicalTrials.gov Identifier: NCT02342795

**1.3. SAP version**

SAP Version 2 – 8/3/2018

**1.4. Protocol version**

Amendment 19 – Version Date: 1/1/2018

**1.5. SAP Revisions**

1.5.1. SAP revision history

- Version 1 – approved 12/1/2014

1.5.2. SAP revision justification

- Version 1 – Original statistical analysis plan and power justification published in the Lopez et al., 2016 and included in the primary site protocol originally approved by the VCU IRB.
- Version 2 – Expanded and amended statistical analysis plan to be more comprehensive in the description of the primary, secondary, and exploratory outcomes and their associated analyses, also included relevant information based on other statistical analysis plan source documents (Gamble et al., 2017; CENIC 2: Project 2 SAP, Version-4/12/2018 provided by E. Donny; Statistical analysis plan sample template for clinical trial disclosure projects, no date available available at: https://www.pfizer.com/files/research/research_clinical_trials/Clinical_Data_Access_Request_Sample_SAP.pdf).

1.5.3. Timing of SAP revision in relation to interim analyses

- Not applicable; no interim analyses

**1.6. Roles and Responsibility**

- Caroline Cobb, PhD[a], drafted and reviewed the SAP
- Miao-Shan Yen, MS[b], drafted and reviewed the SAP
- Le Kang, PhD[b], drafted and reviewed the SAP
- Shumei Sun, PhD[b], advised and reviewed the SAP
- Thomas Eissenberg, PhD[a], advised and reviewed the SAP
- Jonathan Foulds, PhD[c], advised and reviewed the SAP

a. Virginia Commonwealth University, Center for the Study of Tobacco Products, Richmond, VA

b. Virginia Commonwealth University, Department of Biostatistics, Richmond, VA

c. Penn State University Department of Public Health Sciences, Tobacco Center of Regulatory Science, College of Medicine, Hershey, PA

**1.7. Approvals and Date**

Caroline Cobb – 8/8/2018 (via email)

Miao-Shan Yen - 8/8/2018 (via email)

Le Kang - 8/8/2018 (via email)

Shumei Sun - 8/8/2018 (via email)

Thomas Eissenberg - 8/3/2018 (via email)

Jonathan Foulds - 8/3/2018 (via email)

#### Section 2: Introduction

**2.1. Background and rationale**

The tobacco marketplace in the U.S. is changing fast, while regulatory science lags behind. The FDA can alter this dynamic, but needs the tools to do so. One necessary tool is a model for evaluation of all types of "modified risk tobacco products" (MRTPs): novel products marketed with the claim that they reduce harm or risk associated with the use of conventional products. Such claims need to be evaluated and the products regulated accordingly. However, few models have been proposed for this purpose. This project will show how Randomized Controlled Trial (RCT) methods can inform pre-market evaluation by examining the influence of real world product use on biomarkers of toxicant exposure and disease risk, reports of adverse events, and concurrent use of other tobacco products. Electronic cigarettes (ECIGs) are the focus product for this study. Overall, this project will demonstrate how regulatory science is advanced by an integrated, iterative model of MRTP evaluation that includes RCT methods.

**2.2. Objectives**

The specific aims of the RCT are to:

1) *Characterize product influence on toxicants, biomarkers, health indicators, and disease risk.* We will measure exposure to the carcinogenic nitrosamine 4-(methylnitrosamino)-1-(3-pyridyl)-1-butanone [NNK; via its metabolite NNAL (4-(methylnitrosamino)-1-(3-pyridyl)-1-butanol) in urine], expired air carbon monoxide (CO), and nicotine (via its metabolite cotinine in urine). We will also measure heart rate and blood pressure, biochemical and hematologic health indices, pulmonary function (via spirometry), and biomarkers of oxidative stress. With respect to toxicant exposure, we hypothesize that, relative to the cigarette substitute condition, we will observe ECIG liquid nicotine concentration-related decreases in urine NNAL and expired air CO concentration.

2) *Determine the tobacco abstinence symptom and adverse event profile associated with real-world product use*. We will use standard measures of nicotine/tobacco abstinence symptoms (i.e., MNWS, Questionnaire of Smoking Urges) to characterize the extent to which ECIG-induced suppression of abstinence symptoms is related to nicotine

concentration. With respect to other adverse events, we will assess effects likely attributable to inhalation of propylene glycol and nicotine self-administration. We hypothesize more of these propylene glycol-related adverse events with ECIGs relative to the cigarette substitute condition.

3) *Examine the influence of novel product use on conventional tobacco product use.* We will monitor ECIG and all other tobacco/nicotine product use closely, via daily tobacco use diaries and in-person assessments. Because we are targeting individuals interested in reducing their cigarette intake, we hypothesize ECIG nicotine concentration-related reductions in combustible cigarette use.

#### Section 3: Study Methods

##### 3.1. Trial Design

The study is a two-site, four-arm, 6-month, parallel-group randomized controlled trial with a follow-up to 9 months.

##### 3.2. Randomization

Blocked randomization was accomplished with a 1:1:1:1 ratio of condition assignments at each participating institution with the original goal of 260 randomized at each site totaling 520. Based on slower recruitment at the VCU site, this randomization allocation by site was changed (10/5/2015) with VCU randomizing 200 and PSU randomizing 320 (totaling 520). The assignment codes were made from separate randomization lists created in advance by the statistician for each site stratum.

##### 3.3. Sample Size

RCT power was based on the important biomarker of toxicant exposure, NNAL concentration in urine. Unfortunately, there were no data that revealed the NNK exposure in ECIG users, as that study has not yet been performed. However, we do have data showing how NNAL concentration in novel tobacco product users compared to own brand cigarette use (Breland, Kleykamp, & Eissenberg, 2006). Using these data, power analysis revealed 100 completers per condition would provide an 80% power to detect an effect size of 58.59 pg/mL (SD = 125.55 pg/mL) on NNAL. We aimed to enroll 130 participants per condition (across sites) anticipating a 20% attrition rate.

##### 3.4. Framework

3.4.1. Randomized assignment to one of four conditions (cigarette substitute; ECIG with 0 mg/ml nicotine liquid; ECIG with 8 mg/ml nicotine liquid; ECIG with 36 mg/ml nicotine liquid; ECIG nicotine dose administered double-blind).

3.4.2. Primary outcome hypothesis is that relative to the cigarette substitute there will be nicotine dose related decreases in total NNAL.

##### 3.5. Statistical interim analyses and stopping guidance

3.5.1. No interim analyses regarding the primary or secondary outcomes were planned or carried out.

3.5.2. There are no planned adjustments based on interim analyses.

3.5.3. Stopping Rules

3.5.3.1. The Data and Safety Monitor Board (DSMB) is comprised of four scientists not otherwise affiliated with the clinical trial (please see DSMP/DSMB documentation for more information). The DSMB reserves the right to discontinue or suspend the study at any time, at an individual site or overall, for safety or for administrative reasons. In the event of such action, the DSMB will promptly inform the impacted Investigators and institutions, the regulatory authorities, and the Institutional Review Board (IRB) of the action and the reason(s) for the action.

3.5.3.2. Medical Monitors

The roles of the Medical Monitors are to:

- Review and provide definitive adjudication on individual adverse events (AE).
- Make specific recommendations for study product dispensing.

3.5.3.3. Recording and Definition of Adverse Events (per VCU Protocol_A19_1.1.)

Adverse events are recorded and reviewed for updates/changes at every study visit following Visit 1 as well as documented if a participant makes a report via phone or email.

All adverse events (serious or non-serious) and abnormal test findings observed or reported to study team believed to be associated with the study device will be followed until the event (or its sequelae) or the abnormal test finding resolves or stabilizes at a level acceptable to the investigator.

An abnormal test finding will be classified as an *adverse event* if one or more of the following criteria are met:

- The test finding is accompanied by clinical symptoms
- The test finding necessitates additional diagnostic evaluation(s) or medical/surgical intervention; including significant additional concomitant drug treatment or other therapy. **Note:** Simply repeating a test finding, in the absence of any of the other listed criteria, does not constitute an adverse event.
- The test finding leads to a change in study drug dosing or discontinuation of subject participation in the clinical research study.
- The test finding is considered an adverse event by the investigator.

Table 3-1. Adverse Event Definitions

| | |
|---|---|
| **Adverse event** | Any untoward medical occurrence associated with the use of the drug in humans, whether or not considered drug related |
| **Adverse reaction** | Any adverse event caused by a drug |

| | |
|---|---|
| **Suspected adverse reaction** | Any adverse event for which there is a reasonable possibility that the drug caused the adverse event. Suspected adverse reaction implies a lesser degree of certainty about causality than "adverse reaction".<br>• *Reasonable possibility.* For the purpose of IND safety reporting, "reasonable possibility" means there is evidence to suggest a causal relationship between the drug and the adverse event. |
| **Serious adverse event or Serious suspected adverse reaction** | Serious adverse event or Serious suspected adverse reaction: An adverse event or suspected adverse reaction that in the view of either the investigator or sponsor, it results in any of the following outcomes: Death, a life-threatening adverse event, inpatient hospitalization or prolongation of existing hospitalization, a persistent or significant incapacity or substantial disruption of the ability to conduct normal life functions, or a congenital anomaly/birth defect. Important medical events that may not result in death, be life-threatening, or require hospitalization may be considered serious when, based upon appropriate medical judgment, they may jeopardize the patient or subject and may require medical or surgical intervention to prevent one of the outcomes listed in this definition. |
| **Life-threatening adverse event or life-threatening suspected adverse reaction** | An adverse event or suspected adverse reaction is considered "life-threatening" if, in the view of either the Investigator (i.e., the study site principal investigator) or Sponsor, its occurrence places the patient or research subject at immediate risk of death. It does not include an adverse event or suspected adverse reaction that had it occurred in a more severe form, might have caused death. |
| **Unexpected adverse event or Unexpected suspected adverse reaction.** | An adverse event or suspected adverse reaction is considered "unexpected" if it is not listed in the investigator brochure, general investigational plan, clinical protocol, or elsewhere in the current IND application; or is not listed at the specificity or severity that has been previously observed and/or specified. |
| **Unanticipated adverse device effect** | Any serious adverse effect on health or safety or any life-threatening problem or death caused by, or associated with, a device, if that effect, problem, or death was not previously identified in nature, severity, or degree of incidence in the investigational plan or IDE application (including a supplementary plan or application), or any other unanticipated serious problem associated with a device that relates to the rights, safety, or welfare of subjects. |

3.5.3.4 Safety monitoring

The **Principal Investigator** will confirm that all adverse events (AE) are correctly entered into the AE case report forms by the coordinator; be available to answer any questions that the coordinators may have concerning AEs; and will notify the IRB, FDA, sponsor and/or DSMB of all applicable AEs as appropriate. All assessments of AEs will be made by a licensed medical professional who is an investigator on the research.

The **research coordinator** will complete the appropriate report form and logs; assist the PI to prepare reports and notify the IRB, FDA, and/or DSMB of all Unanticipated Problems/SAE's.

Table 3-2. Definitions of Adverse Event Class

| Class | Severity | Expectedness/Relatedness | Location | Reporting Timeline |
|---|---|---|---|---|
| 1 | Serious | - Unexpected<br>- Related or possibly related | All | 2 business days from occurrence |
| 2 | Non-serious | - Unexpected<br>- Related or possibly related | All | Annual |
| 3 | Serious or non-serious | - Expected | All | Annual |

##### 3.6. Timing of outcome assessments

Time points at which each of the primary, secondary, and other/exploratory outcomes are measured including visit "windows" are listed below.

Table 3-3. Timing of Assessments and Study Windows

| | Before Consent | Consent | Reduction Phase | | | | Continuation Phase | | | | | Follow-Up Phase | |
|---|---|---|---|---|---|---|---|---|---|---|---|---|---|
| | | | Baseline | | | | | | | | | | |
| **Study Visit** | **NA** | **1** | **2** | **3** | **4** | **5** | **6** | **7** | **8** | **9** | **10** | **11** | **12** |
| **Study Week** | **NA** | **-1** | **0** | **1** | **2** | **4** | **8** | **12** | **16** | **20** | **24** | **28** | **36** |
| **Days** | **NA** | **-7** | **0** | **7** | **14** | **28** | **56** | **84** | **112** | **140** | **168** | **197** | **253** |
| **Study Visit Window** | **NA** | **-7** | **-1/ +3** | **-3/ +3** | **-3/ +7** | **-7/ +14** | **-7/ +14** | **-7/ +14** | **-7/ +14** | **-7/ +14** | **-7/ +42** | **-7/ +14** | **-7/ +14** |
| **Participant Instructions:** | | | | | | | | | | | | | |
| Smoke normally for one week | | x | | | | | | | | | | | |
| Randomize participants | | | x | | | | | | | | | | |
| Reduce cigarette consumption by 50% | | | x | x | | | | | | | | | |
| Reduce cigarette consumption by 75% | | | | | x | x | | | | | | | |

| | | | | | | | | | | | | | |
|---|---|---|---|---|---|---|---|---|---|---|---|---|---|
| Continue to reduce cigarette consumption | | | | | | | x | x | x | x | | | |
| Cease all combustible tobacco use | | | | | | | | | | | x | | |
| **Baseline Measurements:** | | | | | | | | | | | | | |
| Demographics | | x | | | | | | | | | | | |
| Tobacco Use History | | x | | | | | | | | | | | |
| Treatment History | | x | | | | | | | | | | | |
| Cigarette Details | | x | x | | | | | | | | | | |
| Medical History | | x | | | | | | | | | | | |
| Concomitant Medications | | x | x | x | x | x | x | x | x | x | x | x | x |
| **Drug/Alcohol Measures:** | | | | | | | | | | | | | |
| Alcohol AUDIT-C | | x | | | | x | | | | | x | | x |
| NIDA Quick Screen | | x | | | | | | x | | | x | | x |
| **Adverse events:** | | | | | | | | | | | | | |
| AE Trigger Question | | | x | x | x | x | x | x | x | x | x | x | x |
| AE Log | | | x | x | x | x | x | x | x | x | x | x | x |
| **Cigarette Measures:** | | | | | | | | | | | | | |
| MNWS | | | x | x | x | x | x | x | x | x | x | x | x |
| Confidence to Quit | | | | | | | | | | | x | x | x |
| Stage of Change | | | | | | | | | | | x | x | x |
| Environmental Smoke | | | x | | | | | | | | x | | x |
| Smoking Urges | | | x | x | x | x | x | x | x | x | x | x | x |
| 7-day TLFB & Current Tobacco | | | x | x | x | x | x | x | x | x | x | x | x |
| **Cigarette Dependence** | | | | | | | | | | | | | |
| Cigarette Dependence | | | x | x | x | x | x | x | x | x | x | x | x |
| **Study Product Dependence** | | | | | | | | | | | | | |
| E-Cigarette Dependence Scale (if product = ecig) | | | | x | x | x | x | x | x | x | x | x | x |
| Non-Study E-Cigarette Dependence Scale | | | | x | x | x | x | x | x | x | x | x | x |
| Cig Substitute Dependence Scale (if product = cig sub) | | | | x | x | x | x | x | x | x | x | x | x |

| | | | | | | | | | | | | | |
|---|---|---|---|---|---|---|---|---|---|---|---|---|---|
| E-Cigarette Patterns of Use (if product = ecig) | | | | x | x | x | x | x | x | x | x | x | x |
| **Study Product Measures:** | | | | | | | | | | | | | |
| Study Product Side Effects | | | | x | x | x | x | x | x | x | x | x | x |
| Study Product Evaluation | | | | x | x | x | x | x | x | x | x | x | x |
| Study Product Trigger | | | | x | x | x | x | x | x | x | x | x | x |
| Study Product Dispensed Log | | | x | x | x | x | x | x | x | x | x | | |
| **Psych Measures:** | | | | | | | | | | | | | |
| Kessler 6 | | | x | | | x | x | | x | | x | | x |
| Perceived Stress | | | x | | | x | x | | x | | x | | x |
| CES-D | | | x | | | x | x | | x | | x | | x |
| **Health Measures:** | | | | | | | | | | | | | |
| Interheart | | | x | | | | | | | | x | | x |
| Clinical COPD Questionnaire | | | x | | | | | | | | x | | x |
| **Biomeasures:** | | | | | | | | | | | | | |
| Pulmonary Function Test | | | x | | | x | | x | | | x | | x |
| CO | | x | x | x | x | x | x | x | x | x | x | x | x |
| Waist/Hip Ratio | | | x | | | | | | | | x | | x |
| Vitals (HR/BP) | | x | x | x | x | x | x | x | x | x | x | x | x |
| Height | | | x | | | | | | | | | | |
| Weight | | | x | x | x | x | x | x | x | x | x | x | x |
| **Exhaled Breath Condensate Samples:** | | | x | | | x | | x | | | x | | |
| Oxidative Stress - 8 Isoprostanes & other biomarkers | | | x | | | x | | x | | | x | | |
| **Blood Samples:** | | | | | | | | | | | | | |
| Complete Metabolic Panel | | | x | | | | | | | | x | | |
| Hematology Panel | | | x | | | | | | | | x | | |
| Lipid Panel | | | x | | | | | | | | x | | |
| C-Reactive Protein | | | x | | | | | | | | x | | |
| Oxidative Stress - Glutathione | | | x | | | x | | x | | | x | | |
| **Urine Samples:** | | | | | | | | | | | | | |

| | | | | | | | | | | | | | |
|---|---|---|---|---|---|---|---|---|---|---|---|---|---|
| Pregnancy Test | | x | | | | | | | | | | | |
| Cotinine | | | x | | | x | | x | | | x | | |
| NNAL | | | x | | | x | | x | | | x | | |
| Oxidative Stress - 8 Isoprostanes & 8-OHdG | | | x | | | x | | x | | | x | | |
| **Other:** | | | | | | | | | | | | | |
| Screener 1 | x | | | | | | | | | | | | |
| Screener 2 | x | | | | | | | | | | | | |
| Screener 3 | | x | | | | | | | | | | | |
| End of trial form | | | | | | | | | | | | | X |

### Section 4: Statistical Principles

##### 4.1. Level of statistical significance and adjustment for multiplicity

All statistical tests will be two-sided. The allowed type I error rate is set at 0.05. P-values less than 0.05 will be considered statistically significant, with the exception of the post-hoc analysis of primary outcome between any two research interventions, where p-values less than 0.008 will be considered significant based on Bonferroni corrections for multiple comparison adjustment.

All analyses will be completed using the intent-to-treat (ITT) approach to avoid the bias associated with the non-random loss of participants for the real life intervention effect.

Per-protocol (PP) analysis will also be conducted for the participants who adhered to the study protocol (attended and provided data at Visits 2, 5, 7, and 10) to estimate the true efficacy. Methods for handling missing data will be specified below.

##### 4.2. Confidence intervals to be reported

A 95% Confidence Interval will be reported when appropriate.

##### 4.3. Protocol deviations

4.3.1. Definition of protocol deviations and violations for the trial

**Protocol deviations** are situations where activities of the study diverge from the IRB approved study protocol. They may be accidental or unintentional changes to, or non-compliance with the study protocol that does not increase the risk or decrease the benefit to the participant. It also does not have an effect on the participant's rights, safety, welfare and/or integrity of the data. Deviations may result from the action of the subject, researcher or research staff.

Protocol deviations may not be reportable to the IRB but should be documented in REDCap on the Protocol Deviations Violations form.

**The following will be considered Protocol Deviations throughout the study:**

- **Training:**
    - Study staff who has not been fully trained on study procedures completes visits

- **Researcher oversight:**
  - Study visit offered to a participant that is outside a study window (this does not include a rescheduled visit that is scheduled outside the window due to a conflict in the participant's schedule)
  - Failure on the part of the researcher to collect subject data or specimens (this does not include incidents that are out of the researchers control, i.e. a participant refuses or leaves unexpectedly)
  - Failure to give participants study visit payment
- **Blood/urine samples:**
  - Pregnancy test was not performed on visit dates where required
  - Blood or urine was collected but not documented in REDCap or the specimen logs
  - Blood was not processed and frozen within the 2 hour time window
  - Blood samples were improperly labeled, logged, and/or packaged when shipped but did not result in lost or unidentifiable samples
- **Clinic procedures:**
  - Incorrect information was entered into spirometry software yielding incorrect predicted values
- **Participants:**
  - Subject refuses to complete research activities that they have otherwise consented to (questionnaires, blood/urine collection etc.)
- **Confidentiality:**
  - Inclusion of identifying information on sample shipments
- **Study Product:**
  - Cartomizers given to participant after expiration date

**Protocol violations** are more serious and may reduce the quality or completeness of the data, make the informed consent inaccurate or impact the participant's safety, rights or welfare.

Protocol violations **are reportable to the PSU IRB** and if they result in an **unanticipated problem are reportable to the VCU IRB**.

**The following will be considered protocol violations throughout the trial:**

- **Confidentiality:**
  - Breaches of confidentiality resulting from lost, misplaced, or stolen study documents (e.g., a consent form is missing)
  - Inclusion of identifying information on sample shipments
- **Consent form:**
  - Failure to obtain valid informed consent prior to any study-specific tests/procedures
  - Missing signature or date from either participant or researcher
  - Outdated or incorrect consent form used
- **Randomization:**
  - A participant is consented into the study who does not meet initial inclusion/exclusion criteria
  - Participant is entered into randomization phase prior to determining final compliance criteria
- **IRB:**
  - Unreported SAEs to the IRB within 5 days
  - Approvals not current, suspended or terminated

  - Enrollment occurs during a period when study is “on hold”
  - Enrollment over the IRB approved enrollment total
- **Tests/Samples:**
  - Incorrect or missing tests (PFT, blood, EBC or urine samples) that were documented as complete
  - Mishandled samples: Blood, urine, or EBC samples were not labeled, logged, and/or packaged properly when shipped resulting in lost samples
- **Safety:**
  - Participant was allowed to continue with the study after a positive pregnancy test
  - Participant was given study product when withdrawal criteria was met
  - Participant was allowed to continue with the study after any other hard withdrawal criteria are met
- **Participants:**
  - Visits are repeatedly scheduled outside study windows
- **Data Collection:**
  - Phone and/or survey contacts were not attempted as scheduled due to researcher oversight

4.3.2. Description of which protocol deviations will be summarized

Percentages of protocol deviations related to research oversight, blood/urine samples, clinic procedures, and study products will be summarized by site and by conditions.

##### 4.4. Analysis populations

4.4.1. The primary analysis of all endpoints will adhere to the ITT principle, while PP analysis will also be considered to provide additional insights about true efficacy. Under the ITT principle, all randomized subjects will be included in the analysis in the group to which they were randomized, regardless of protocol violations and compliance to treatment assignment; while the PP analysis will be restricted to those participants who adhered to the study protocol.

###### 4.4.2. Subgroup populations

None are planned.

#### Section 5: Trial Population

##### 5.1. Screening Data and Eligibility

Participant screening data (collected via phone via a pre-screener and in-person at Visit 1) will be reported for all items related to primary inclusion and exclusion criteria. Inclusion and exclusion eligibility criteria are described below.

Inclusion Criteria

- Age 21-65
- Smoke >9 cigarettes per day for at least 1 year
- Smoke regular filtered cigarettes or machine-rolled cigarettes with a filter
- CO measurement >9 ppm at baseline
- No serious quit attempt in the prior 1 month. This includes use of any FDA approved smoking cessation medication (varenicline, bupropion [used specifically as a quitting

aid], patch, gum, lozenge, inhaler, and nasal spray) in the past 1 month as an indication of treatment seeking.
- Not planning to quit in the next 6 months
- Interested in reducing cigarette consumption
- Willing to attend visits weekly and monthly over a 9-month period (not planning to move, not planning extended vacation, no planned surgeries
- Read and write in English
- Able to understand and consent

Exclusion Criteria

- Pregnant and/or nursing women
- Unstable or significant medical condition in the past 12 months (Recent heart attack or some other heart conditions, stroke, severe angina including high blood pressure if systolic >159 or diastolic >99 observed during screening).
- Severe immune system disorders (uncontrolled HIV/AIDS; unstable multiple sclerosis symptoms), respiratory diseases (exacerbations of asthma or COPD, require oxygen, require oral prednisone), kidney (dialysis) or liver diseases (cirrhosis), or any medical disorder/medication that may affect participant safety or biomarker data.
- Use of any non-cigarette nicotine delivery product (pipe, cigar, dip, chew, snus, hookah, ECIGs, strips, sticks) in the past 7 days
- Uncontrolled mental illness or substance abuse or inpatient treatment for these in the past 6 months
- History of difficulty providing or unwilling to provide blood samples (fainting, poor veins, anxiety)
- Surgery requiring general anesthesia in the past 6 weeks
- Use of an ECIG for 5 or more days in the past 28 days or any use in the past 7 days
- Use of marijuana or any illicit drug/prescription drugs for non-medical use daily/almost daily or weekly in the past 3 months per NIDA Quick Screen
- Use of hand-rolled, roll your own cigarettes
- Known allergy to propylene glycol or vegetable glycerin
- Other member of household participated in the study

##### 5.2. Recruitment

The CONSORT flow diagram for the study will include the proportion of the sample assessed for eligibility via the pre-screener, eligible/ineligible for Visit 1, attended/excluded/eligible at Visit 1, attended/excluded/randomized at Visit 2, study condition/arm assigned at Visit 2, and for Visits 3-12 attended/no-show/withdrawn (reason for withdrawal) by study condition/arm assigned.

##### 5.3. Withdrawal/follow-up

Withdrawal from the study will be defined using the end of trial form (PI decision, participant decision to withdraw, withdrawal for serious adverse event). Sub-categories for each type of withdrawal will be presented by study condition assigned and by site.

##### 5.4. Baseline participant characteristics

Baseline characteristics to be summarized include demographics (age, sex, race, ethnicity, education, employment status, total household income, marital status), tobacco use characteristics (average cigarettes smoked per day, years smoking, age of cigarette smoking initiation, ever use of other tobacco products, menthol status, PSU Cigarette Dependence Scale score, Visit 2 average cigarettes per day), and psychosocial/health characteristics (AUDIT score, Environmental Smoke Score, Kessler K6 Score, Perceived Stress Score, CES-D Score, Interheart Score, Clinical COPD Score), physiological measures (waist circum, hip circum, heath, weight, BMI, BP, Pulse, FEV1, FVC, urine cotinine, urine NNAL, and expired air CO).

#### Section 6: Analysis

##### 6.1. Outcome definitions and timing

- Primary outcome: The primary outcome measure of this RCT is the urinary concentration of the carcinogen biomarker of tobacco exposure, total NNAL (linear range: 20 to 20,000 pg/mL or 0.020 to 20 ng/mL. Results will be presented unadjusted and adjusted for creatinine concentration (pg/ng per mg creatinine). This outcome is measured at Visit 2 (Week 0), Visit 5 (Week 4), Visit 7 (Week 12), and Visit 10 (Week 24).

- Secondary outcomes:
  ○ Cotinine concentration will be measured via urine sample (linear range: 1000 to 3,000,000 pg/mL or ng/mL or 1-3000 ng/mL). Results will be presented unadjusted and adjusted for creatinine concentration (pg/ng per mg creatinine). This outcome is measured at Visit 2 (Week 0), Visit 5 (Week 4), Visit 7 (Week 12), and Visit 10 (Week 24).
  ○ Total glutathione will be measured via blood sample (linear range information unavailable). This outcome is measured at Visit 2 (Week 0), Visit 5 (Week 4), Visit 7 (Week 12), and Visit 10 (Week 24).
  o 8-Isoprostanes will be measured via urine sample (linear range information unavailable). Results will be presented unadjusted and adjusted for creatinine concentration (pg/ng per mg creatinine. This outcome is measured at Visit 2 (Week 0), Visit 5 (Week 4), Visit 7 (Week 12), and Visit 10 (Week 24).
  ○ 8-Isoprostanes will be measured via exhaled breath condensate sample (linear range information unavailable). This outcome is measured at Visit 2 (Week 0), Visit 5 (Week 4), Visit 7 (Week 12), and Visit 10 (Week 24).

- Exploratory outcomes (timing varies between Visit 1-12):
  - From Aim 1
    - Exhaled CO
    - Heart rate/blood pressure
    - Spirometry outcomes
    - 8-OHdG in urine samples
    - Other oxidative stress markers in exhaled breath condensate samples.
    - Blood lab results
  - From Aim 2
    - Adverse events profile
      - By overall frequency, by relatedness category, severity, and by system affected
    - Study Product Side Effects
    - Study Product Evaluation

            - Minnesota Nicotine Withdrawal Scale (MNWS)
            - Questionnaire of Smoking Urges
    - From Aim 3
        - 7-day TLFB form
            - Cigarettes smoked per day (7-day average)
            - Study product use per day (7-day average)
            - Study product cartomizers dispensed/used returned
            - 50% reduction in cigarettes smoked (defined as 50% reduction of Visit 1 self-reported CPD; yes/no)
            - 75% reduction in cigarettes smoked (defined as 50% reduction of Visit 1 self-reported CPD; yes/no)
            - Study product abstinence
                - 24-hour use (available at Visits 3-12; yes/no)
            - Cigarette smoking abstinence
                - 24-hour point prevalence cigarette smoking abstinence (available at Visits 3-12; yes/no)
                - 7-day point prevalence cigarette smoking abstinence (available at Visits 3-12; yes/no)
                - 28-day point prevalence cigarette smoking abstinence (available at Visits 6-11; yes/no)
            - Other tobacco use
                - For each product reported 7-day average
                - Summary across products 7-day average
        - Cigarette Dependence Scale
        - Non-Study E-Cigarette Dependence Scale
        - Cigarette Substitute Dependence Scale
        - E-Cigarette Dependence Scale
        - E-Cigarette Patterns of Use
        - Study Product Dispensed Log
    - Other outcomes
        - AUDIT-C
        - CES-D
        - Clinical COPD Questionnaire
        - Perceived Stress
        - Kessler K6
        - Interheart
        - Environmental Smoke Questionnaire

6.1.2. Any calculation or transformation used to derive the primary and secondary outcomes (i.e., change from baseline)

None planned other than corrected/non-corrected values for creatinine where applicable and potential transformation for outcomes that do not meet model assumptions such as normality.

6.1.3. Any calculation or transformation used to derive the exploratory outcomes (i.e., change from baseline)

Changes from baseline may be calculated for some exploratory analyses as needed. Sum scores of MPSS, FTND, PSCDI, HONC, AUDIT-C, MNWS-R, Perceived Stress, Kessler 6, CES-D, Interheart and Clinical COPD will be calculated. Level of exposure to environmental smoke will be categorized based on the responses from the environmental smoke questionnaire. Averages of 7-CPD/study product use will be calculated based on the 7-day TLFB form (please note days that overlap in the 7-day TLFB will be dropped from the more recent visit).

##### 6.2. Analysis methods

6.2.1. Primary and Secondary Analyses

We will first examine baseline demographics described above to identify any baseline imbalances after randomization. Discrete variables will be summarized by frequencies with percentages and compared using Chi-squared test or Fisher's exact test when appropriate. Continuous covariates will be summarized by mean with standard deviation, or median with range, and compared using one-way ANOVA or Kruskal-Wallis test. We expect groups to be balanced for important baseline demographics due to the nature of RCT.

Our primary endpoint, urine NNAL concentration, will be summarized by study intervention and timepoint and analyzed using two-way ANOVA with repeated measures (between-subjects factor: study condition/arm; within-subjects factor: visit, 2, 5, 7, 10), and we will adjust the type I error rate to account for 6 pairwise comparisons between study conditions/arms over time, and at each time point. We will compare each ECIG group to the cigarette substitute and then each ECIG group to the other. An analogous approach will be used to analyze our secondary endpoints. The analyses of our secondary endpoints will primarily use one-way ANOVA at different timepoint with similar multiple comparison adjustments.

Secondary data analyses will consist of adjusted analyses including age, gender, race, and possibly for other covariate imbalance (including by site) with the use of linear mixed models.

6.2.2. Any adjustment for covariates

A secondary analysis (as described above) will be completed adjusting for age, gender, race, along with any other covariates that significantly differ across study conditions at baseline, or any other covariates particularly relevant to the primary/secondary outcomes. A stepwise model selection will be performed to finalize the list of covariates to be included in the final model.

6.2.3. Methods used for assumptions to be checked for statistical methods

Data normality will be confirmed by using graphical methods including quantile-quantile plot (QQ plot) and Kolmogorov-Smirnov test.

6.2.4. Details of alternative methods to be used if distributional assumptions do not hold

Skewed continuous variables will be log-transformed or Box-Cox transformed as appropriate.

##### 6.3. Safety

The distinction between the primary/secondary outcomes and safety outcomes is not as clear in this trial as it would be in a typical clinical trial of a novel therapeutic agent. Many outcomes that would typically be considered potential adverse consequences or safety outcomes will be analyzed as the secondary or exploratory outcomes as described above.

AEs and SAEs will be recorded as described in the Adverse Event SOP. AEs will be tabulated and compared across treatment groups in the exploratory analyses. We expect study product-related SAEs to be rare and, therefore, no formal statistical comparison for this AE category is planned for this trial.

##### 6.4. Missing data

An ITT analysis includes subjects that are allocated according to their random intervention assignment, regardless of compliance to the study product, and that complete data are available on all study subjects. Every effort will be made to limit the amount of missing data in this trial. Study participants will be incentivized to attend study sessions and provide biological samples and other measurements as detailed in the study protocol. However, some level of missing data is inevitable in a study of this kind. In response, we will complete a sensitivity analysis for the primary and secondary endpoints in order to evaluate the robustness of our conclusions to missing data.

We will compare subjects with and without missing primary outcome data in order to identify baseline covariates associated with missing data. There are three possible scenarios of missing data in our study: (1) primary outcome measurements missing while covariate measurements completely available, (2) primary outcome measurements available but some measurements in covariates missing, and (3) both primary outcome and some covariate measurements missing. Our primary approach to handling missing data in scenario 1 and 2 will be multiple imputation where missing values are imputed using conditional regression models developed from baseline covariates (Little & Rubin, 2002) and/or available outcomes. Our strategy to handle missing data in scenario 3 is multiple imputation through joint conditional specification (Schafer, 1997).

In addition, we will conduct a series of sensitivity analyses of the primary and secondary endpoints using baseline-carried-forward and last-observation carried forward. The results of these analyses will be compared to the primary analysis to evaluate the robustness of our conclusions.

Note that we intent to examine the potential differential dropout or attrition between interventions and study sites. If there is information suggesting data missing are not at random, e.g., the dropout rates are evidently different between interventions or sites, multiple imputation model equations should not incorporate intervention or site variable. Alternatively, multiple imputation may be carried out one intervention at a time, or one site at a time, respectively (Li, Stuart, & Allison, 2015).

##### 6.5. Additional analyses

Exploratory endpoints will be analyzed following a similar approach to what has been proposed for the primary and secondary endpoints. Continuous and binary endpoints will be analyzed using an ANOVA, linear regression, or logistic regression, respectively, and secondary data analyses will be performed using linear mixed models or generalized linear mixed models, as appropriate. Skewed continuous endpoints will be log-transformed and analyzed on the log-scale for analysis. Time-to-event endpoints will be analyzed using Cox proportional hazards model. The primary analysis of all endpoints will be without adjustment for baseline covariates.

The secondary analysis will be completed adjusting for age, gender, and race, along with any other covariates that differ significantly across study conditions at baseline, or any other covariates particularly relevant to the exploratory outcomes (including by site). A stepwise model selection will be performed to finalize the covariates to be included in the final model.

Other exploratory analyses include:

1) An exploratory analysis of the primary, secondary, and exploratory endpoints will involve recoding of the study condition assignment into a three-level variable: cigarette substitute s. ECIG+0 mg/ml nicotine vs. ECIG+8/36 mg/ml nicotine (i.e., the two nicotine containing conditions will be collapsed).

2) An exploratory analysis of the primary, secondary, and exploratory endpoints will involve adding the use of other tobacco products (combustible/non-combustible) as a covariate (assessed at each study visit).

3) An exploratory analysis of the primary, secondary, and exploratory endpoints will involve adding the current cigarette smoking and/or cigarette dependence scale score as a covariate (assessed at each study visit).

4) An exploratory analysis of the primary, secondary, and exploratory endpoints by site (VCU vs. PSU) due to anticipated demographic heterogeneity in the population recruited.

5) An exploratory analysis of the primary, secondary, and exploratory endpoints by study product adherence status (i.e., use of study product at subsequent visits following randomization).

##### 6.6. Statistical software

All analysis will be completed using SAS (v.9.4).

### Appendix

**SAP – Version 1**

---

Randomized Control Trial Methods for Novel Tobacco Product Evaluations

Statistical Analysis Plan

Version 1

Authors: Miao-Shan Yen, ALL authors from paper? +_Bullen?

---

Per the Lopez et al. BMC Public Health (2016) 16:217 DOI 10.1186/s12889-016-2792-8

Aim

The specific aims of the RCT are to:

1. Characterize ECIG influence on toxicants, biomarkers, health indicators, and disease risk. We will measure exposure to the carcinogenic nitrosamine 4-(methylnitrosamino)-1-(3-pyridyl)-1-butanone [NNK; via its metabolite NNAL (4-(methylnitrosamino)-1-(3-pyridyl)-1-butanol) in urine], expired air carbon monoxide (CO), and nicotine (via its metabolite cotinine in urine). We will also measure heart rate and blood pressure, biochemical and hematologic health indices, pulmonary function (via spirometry), and biomarkers of oxidative stress. With respect to toxicant exposure, we hypothesize that, relative to the cigarette substitute condition, we will observe ECIG liquid nicotine concentration-related decreases in urine NNAL and expired air CO concentration.
2. Determine the tobacco abstinence symptom and adverse event profile associated with real-world ECIG use. We will use standard measures of nicotine/ tobacco abstinence symptoms to characterize the extent to which ECIG-induced suppression of abstinence symptoms is related to nicotine concentration. With respect to other adverse events, we will assess effects likely attributable to inhalation of propylene glycol and nicotine self administration.
3. We hypothesize more of these propylene glycol-related adverse events with ECIGs relative to the cigarette substitute condition. Examine the influence of ECIG use on conventional tobacco product use. We will monitor ECIG and all other tobacco/nicotine product use closely, via daily tobacco use diaries and in-person assessments. Because we are targeting individuals interested in reducing their cigarette intake, we hypothesize that individuals randomized to higher ECIG nicotine concentrations (8 mg/ml or 36 mg/ml) will experience reductions in combustible cigarette use.

Primary outcome

The primary outcome measure of this RCT is the urinary concentration of the carcinogen biomarker of tobacco exposure, NNAL.

Secondary outcomes

The secondary outcome measure will be urine cotinine concentration. Markers of oxidative stress (Glutithione and 8-Isoprostanes) are additional secondary outcome measures. Glutathione will be measured via blood sample analysis. 8-Isoprostanes will be measured via both urine sample and exhaled breath condensate analyses.

Sample size

RCT power is based on the important biomarker of toxicant exposure, NNAL concentration in urine. Unfortunately, there are no data that reveal the NNK exposure in ECIG users, as that study has not yet been performed. However, we do have data showing how NNAL concentration in novel tobacco product users compares to own brand cigarette use [49]. Using these data, power analysis revealed 100 completers per condition would provide an 80 % power to detect an effect size of 0.28 pmol/ml (SD = 0.6 pmol/ml) on NNAL. We aim to enroll 130 participants per condition anticipating a 20 % attrition rate.

Statistical analysis

The analysis plan is based on the primary objective of determining the extent to which ECIG nicotine concentration influences NNK exposure as indexed by urinary NNAL concentration. We will first examine baseline characteristics including demographics and smoking history across study interventions to identify any baseline imbalances after randomization. Discrete variables will be summarized by frequencies and percentages and compared using Chisquared test or Fisher's exact test. Continuous covariates will be summarized by mean, standard deviation, median and range, and compared by one-way ANOVA. Skewed continuous variables will be log-transformed or square root transformed as appropriate. We expect groups to be balanced for important baseline characteristics due to randomization.

A secondary analysis will be completed adjusting for age, gender, and race, along with any other covariates that differ across research interventions at baseline with a p-value less than 0.20. P-values less than 0.05 will be considered statistically significant with the exception of analysis of our primary analysis, where p-values less than 0.008 will be considered significant after a Bonferroni multiple comparisons adjustment.

Our primary endpoint, urine NNAL concentration, will be summarized by study intervention and time and analyzed using linear regression, and we will adjust the Type I error rate to account for 6 pairwise comparisons at each time point. We will compare each ECIG group to the cigarette substitute and then each ECIG group to the other. An analogous approach will be used to analyze our secondary endpoints. The primary analysis of our secondary endpoints will use linear regression.

Secondary analyses will consist of an adjusted analysis and a repeated measures analysis using a linear mixed model. All analysis will be completed using SAS (v.9.4) under the expertise of a senior biostatistician.

PER VCU approved protocol (dated 10/29/2014; approved by IRB 12/01/2014)

Statistical Plan

7.0 Sample size determination

We chose to power our RCT based on the important biomarker of toxicant exposure, NNAL concentration in urine. Unfortunately, there are no data that reveal the NNK exposure in electronic cigarette users, as that study has not yet been performed. However, we do have data showing how NNAL concentration in novel tobacco users compares to own brand use. Using these data, and the knowledge that we would be conducting two sample t-tests at the .008 level to control for 6 pairwise comparisons we worked with the biostatistics team to conduct a power analysis. That analysis revealed that 100 completers per condition would provide an 80% power to detect an effect size of .28. We have chosen therefore to enroll 130 participants in each condition, in anticipation of a ~20% attrition rate (Note: We will limit attrition by replacing participants before randomization).

#### 7.1 Statistical methods

Our analysis plan is based on the primary objective of determining the extent to which ECIG dose influences NNK exposure as indexed by urine NNAL level. We will first examine baseline characteristics including demographics and smoking history across research interventions to identify any baseline imbalances after randomization. Discrete variables will be summarized by frequencies and percentages and compared using Chi-squared test of Fisher's exact test. Continuous covariates will be summarized by mean, standard deviation, median and range, and compared by one-way ANOVA. Skewed continuous variables will be log-transformed or square root transformed as appropriate. We expect groups to be balanced for important baseline characteristics due to randomization. A secondary analysis will be completed adjusting for age, gender, and race, along with any other covariates that differ across research interventions at baseline with a p-value less than .20. P-values less than .05 will be considered statistically significant with the exception of analysis of our primary analysis, where p-values less than .008 will be considered significant after a Bonferroni multiple comparisons adjustment. Our primary endpoint, urine NNAL level, will be summarized by research intervention and time and analyzed using linear regression and we will adjust the Type 1 error rate to account for 6 pairwise comparisons at each time point. We will compare each electronic cigarette group to the control and then each electronic cigarette group to the other. An analogous approach will be used to analyze our secondary endpoints. The primary analysis of our secondary endpoints will use linear regression. Secondary analyses will consist of an adjusted analysis and a repeated measures analysis using a linear mixed model. All analysis will be completed using SAS (v.9.3).
